# Long-term ambient hydrocarbon exposure and incidence of urinary bladder cancer

**DOI:** 10.1101/2022.05.08.22274798

**Authors:** Han-Wei Zhang, Zhi-Ren Tsai, Victor C Kok, Hsiao-Ching Peng, Yau-Hung Chen, Jeffrey JP Tsai, Chun-Yi Hsu

**Affiliations:** Program for Aging, China Medical University, Taichung, Taiwan; Institute of Population Health Sciences, National Health Research Institutes, Miaoli, Taiwan; Institute of Electrical Control Engineering, Department of Electrical and Computer Engineering, National Yang Ming Chiao Tung University, Hsinchu, Taiwan; Biomedica Corporation, New Taipei, Taiwan; Department of Computer Science and Information Engineering, Asia University, Taichung, Taiwan; Department of Medical Research, China Medical University Hospital, China Medical University, Taichung, Taiwan; Center for Precision Medicine Research, Asia University, Taichung, Taiwan; Department of Bioinformatics and Medical Engineering, Asia University, Taichung, Taiwan; Division of Medical Oncology, Kuang Tien General Hospital Cancer Center, Taichung, Taiwan; Professor, Department of Chemistry, Tamkang University, New Taipei City 25137, Taiwan; Graduate Institute of Biomedical Science, China Medical University, Taichung, Taiwan

**Author notes:** **Corresponding Author:** Victor C Kok, MD, MMSc, PhD, FACP, Division of Medical Oncology, Kuang Tien General Hospital Cancer Center, Taichung, Taiwan. Department of Bioinformatics and Medical Engineering, Asia University Taiwan, Taichung, Taiwan.,;, Postal address: 117 Shatien Rd Shalu Dist., Taichung 43303 Taiwan. These two authors share first authorship (Han-Wei Zhang, Zhi-Ren Tsai).

**Keywords:** air pollution, total hydrocarbons, nonmethane hydrocarbon, cancer risk, exposure, xenobiotics, bladder cancer

## Abstract

**Background:** Particulate matter and volatile organic compounds, including total hydrocarbons (THCs), are major ambient air pollutants. The primary nonmethane hydrocarbons (NMHCs) originate from vehicle emissions. Studies on the association between air pollution and urinary bladder cancer (UBC) have revealed contradictory results.

**Objectives:** The present study investigated whether long-term exposure to ambient hydrocarbons increases UBC risk among people aged ≥20 years in Taiwan.

**Methods:** Linkage dataset research with longitudinal design was conducted on 600,666 cancer-free individuals from 2000–2013; 12 airborne pollutants were determined. Several Cox models considering potential confounders were employed. The study outcomes were invasive or *in situ* UBC incidence over time. The targeted pollutant concentration was divided into three tertiles: T1/T2/T3.

**Results:** The mean age of the individuals at risk was 42.8 (SD, 15.7), and 50.2% were men. The mean daily average over 10-years of airborne THC concentration was 2.24 ppm (SD, 0.14), and NMHC was 0.29 ppm (SD, 0.09). There was a dose-dependent increase in UBC at follow-up. The incidence of UBC cases per 100,000 people by T1/T2/T3 exposure to THC was 60.3, 203.7, and 450.8, respectively; it was 180.2/202.4/453.8 per 100,000, corresponding to T1/T2/T3 exposure to NMHC, respectively. Without controlling for confounding air pollutants, the adjusted hazard ratio (adj.HR) was 1.77 (95% CI, 1.69–1.87) per 0.14 ppm increase of THC; after controlling for PM_2.5_, adj.HR was even higher at 2.10 (95% CI, 1.99– 2.22). The adj.HR was 1.38 (95% CI, 1.31–1.44) per 0.09 ppm increase in ambient NMHC concentration. After controlling for SO_2_ and CH_4_, the adj.HR was 1.16 (95% CI, 1.11–1.21). Sensitivity analyses showed that the UBC development risk was not sex-specific or influenced by diabetes status.

**Discussion:** Long-term exposure to THC and NMHC may be a risk factor for UBC development. Acknowledging the pollutant sources can inform risk management strategies.

## Introduction

Anthropogenic environmental pollutants are believed to account for a sizable portion of the worldwide incidence of cancer ^1^. Over the past decades, hundreds of confirmed and suspected environmental carcinogens have been identified. Humans cannot live well without clean air. However, industrialization has led to air pollution from dust storms, smoke, fumes, and toxic gas emissions from thermal power plants, coal mines, petroleum, and chemicals. As a result, ambient air pollution has become one of the most significant environmental risks to health. In addition, exposure to outdoor air pollution poses an urgent public health challenge worldwide, because it is ubiquitous, affects everyone, and has numerous adverse human health effects, including cancer ^2,3^. For example, diesel exhaust and particulate matter (PM) air pollution have been associated with an increased risk of lung cancer ^4-7^. According to the International Agency for Research on Cancer (IARC), air pollution is a Group 1 carcinogen; in addition to causing lung cancer, it is also associated with an increased risk of other types of cancer ^8^. Therefore, studies on the toxicological effects of these anthropogenic ambient air pollutants and their impact on human organs are urgently needed.

Typical sources of air pollutants include fossil fuel combustion, diesel vehicle emissions, smelters, wildfires, biomass burning, gas-to-particle conversion, dust storms, windblown soil, petrochemical solvents, evaporated fuels, incomplete combustion, chemical processing, biogenics, and photochemical reactions in the atmosphere ^1^. Examining the pollutant profiles can inform policy-making on forming environmental and public health counteractive measures; exhaust emissions from transportation include total hydrocarbons (THC) and nitrogen oxides (NO_X_); exhaust gas emitted by industrial plants contain a variety of pollutants, such as volatile organic compounds (VOCs), nitrogen dioxide (NO_2_), and carbon monoxide (CO) ^1^.

Outdoors, petrochemical solvent spillage, evaporated fuels, and biogenic emissions produce air pollutants in the form of gases, VOCs, and PM. Although regarded as low-carbon emissions, electricity generation from natural gas power plants produces airborne nonmethane hydrocarbons (NMHC) and nitrogen oxides (NOx) in the long run. Notably, after a series of photochemical reactions between VOCs (including hydrocarbons) and NO_X_, the concentration of ground-level ozone (O_3_) increases, leading to poor air quality ^9^. Therefore, effective control of hydrocarbons can indirectly reduce the ground-level O_3_ concentration to improve regional air quality.

VOCs are defined in several ways. Total hydrocarbons (THCs) are also VOCs when used in a broader sense. Oxygenated hydrocarbons, such as alcohols and aldehydes, are not considered THCs. Hydrocarbons are the most toxic organic gases in vehicle emissions. Methane, a hydrocarbon, is neither photoreactive nor toxic; conversely, “nonmethane hydrocarbons” are known to be reactive in ambient air. Although slight differences may occur among different geographical regions ^9^, a source apportionment study of non-biogenic, anthropogenic NMHC as an air pollutant in Delhi, India, disclosed that the primary source of NMHC was traffic vehicle emissions (petrol and diesel), with 38% from petroleum, 32% from liquefied petroleum gas, 16% from solid fuel combustion, and 14% from diesel ^10^.

The global cancer burden estimation report, GLOBOCAN 2020, was published in 2021, showing urinary bladder cancer (UBC) as the 12^th^ most common cancer worldwide, accounting for 3% of all cancer burden ^11^. In 2020, there were 573,278 newly diagnosed UBCs, with 212,536 new deaths the same year ^11^. Because UBC is not easily detected early, the mortality rate is relatively high. Furthermore, although the overall number of UBC cases is relatively minor compared with lung cancer, colorectal cancer, and liver cancer, the recurrence rate of bladder cancer is high, by as much as 70%. Fortunately, over the years, through government-led air pollution control, workplace health promotion, industrial occupational hazard exposure prevention, ban on aristolochic acid-containing medicines, and a substantial reduction in the cigarette smoking rate in Taiwan, the age-specific incidence rates of UBC in Taiwan are expected to decrease by >25% from 2016 to 2025 ^12-14^. It is important to examine the magnitude of UBC development risk due to specific ambient air pollutants.

Studies on the association between air pollution and UBC have revealed contradictory results. A few studies with positive results suggest that air pollution, particularly PM, is associated with a higher incidence of UBC ^5,6,15-20^. In contrast, other studies did not detect the causal relationship ^7,21-23^. People living near chemical factories ^24^ and road transportation workers ^6^ harbor a higher risk of developing UBC. However, the culprit pollutants are thought to be airborne polycyclic aromatic hydrocarbons (PAHs) and motor vehicle engine exhaust. In addition, several studies have suggested that air pollution (ambient PM_2.5_, traffic air pollution, and petrochemical air pollutant emissions) is associated with increased bladder cancer mortality in Taiwan ^17,19,21^. However, to our knowledge, no studies have investigated the ambient air hydrocarbon pollutant, THC, and NMHC.

We hypothesized that exposure to the ambient air pollutants THC and NMHC would increase the risk of developing UBC. Data from the National Health Insurance Research Database (NHIRD) and government environmental databases were used to examine whether long-term exposure to hydrocarbons in ambient air increased UBC risk among people aged ≥20 years in Taiwan. This is one of the first studies to investigate the risk of UBC associated with exposure to ambient air THC and NMHC pollution.

## Results

### Study population characteristics

This study tracked 600,666 cancer-free individuals aged 20 years and above (Fig. 1). The mean age was 42.8 ± 15.7. Males accounted for 50.2% of the total population. The most prevalent medical comorbidities were dyslipidemia (23.3%), hypertension (22.8%), diabetes mellitus (17%), chronic liver disease (12.5%), and gout (10.9%). We also examined risk factors for UBC in the entire cohort, which included smoking-related diagnoses with a prevalence of 6.3%, alcohol use disorder (2.1%), morbid obesity (0.9%), spinal cord injury (0.7%), chronic cystitis (0.3%), chronic kidney disease (3.1%), and pesticide exposure (0.1%). The demographic data and comorbid states among tertiles of THC and NMHC are presented in Tables 1 and 2, respectively, with T1 and T3 being the lowest and highest levels of the daily average of the respective pollutant. Individuals exposed to any pollutant in T1 had more comorbidities and risk factors than those under T3 exposure (Table 1 and Table 2).

**Table 1.**
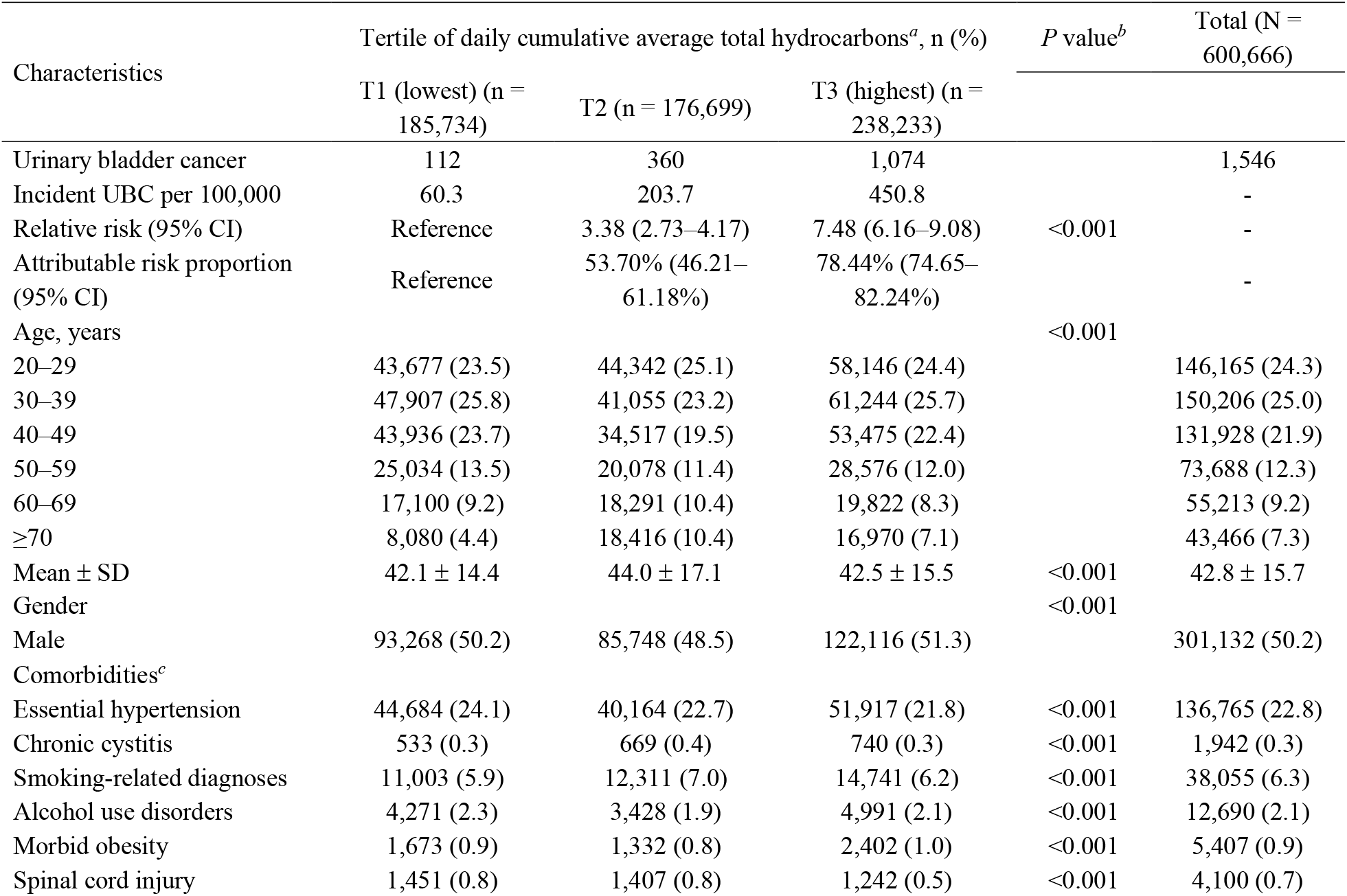
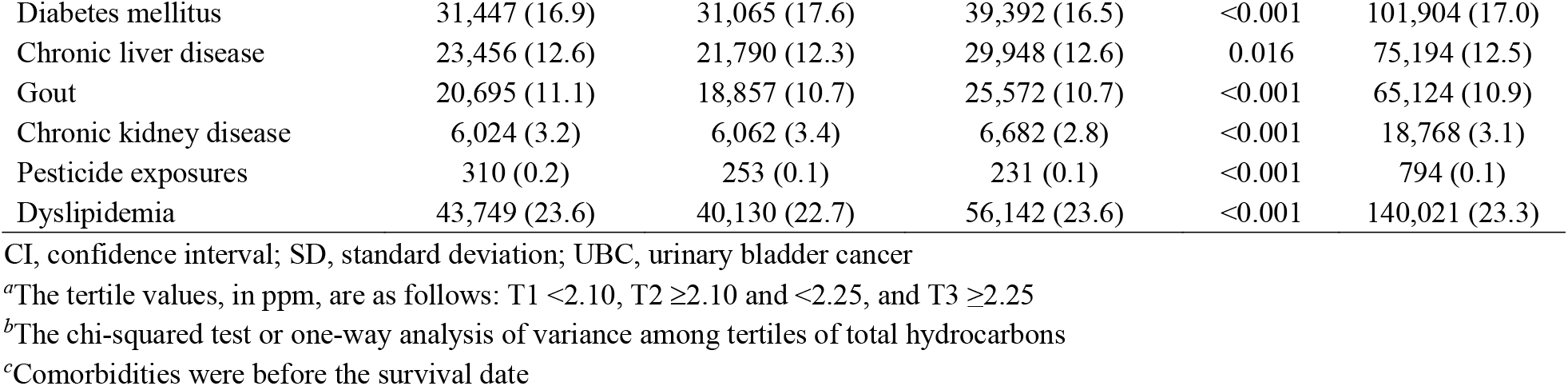
**Relative risks of incident UBC by tertile of the total hydrocarbons’ exposure and characteristics of the cohorts**

| Characteristics | Tertile of daily cumulative average total hydrocarbons<sup>a</sup>, n (%) |  |  | *P* value<sup>b</sup> | Total (N = 600,666) |
| --- | --- | --- | --- | --- | --- |
|  | T1 (lowest) (n = 185,734) | T2 (n = 176,699) | T3 (highest) (n = 238,233) |  |  |
| Urinary bladder cancer | 112 | 360 | 1,074 |  | 1,546 |
| Incident UBC per 100,000 | 60.3 | 203.7 | 450.8 |  | - |
| Relative risk (95% CI) | Reference | 3.38 (2.73–4.17) | 7.48 (6.16–9.08) | <0.001 | - |
| Attributable risk proportion (95% CI) | Reference | 53.70% (46.21–61.18%) | 78.44% (74.65–82.24%) |  | - |
| Age, years |  |  |  | <0.001 |  |
| 20–29 | 43,677 (23.5) | 44,342 (25.1) | 58,146 (24.4) |  | 146,165 (24.3) |
| 30–39 | 47,907 (25.8) | 41,055 (23.2) | 61,244 (25.7) |  | 150,206 (25.0) |
| 40–49 | 43,936 (23.7) | 34,517 (19.5) | 53,475 (22.4) |  | 131,928 (21.9) |
| 50–59 | 25,034 (13.5) | 20,078 (11.4) | 28,576 (12.0) |  | 73,688 (12.3) |
| 60–69 | 17,100 (9.2) | 18,291 (10.4) | 19,822 (8.3) |  | 55,213 (9.2) |
| ≥70 | 8,080 (4.4) | 18,416 (10.4) | 16,970 (7.1) |  | 43,466 (7.3) |
| Mean ± SD | 42.1 ± 14.4 | 44.0 ± 17.1 | 42.5 ± 15.5 | <0.001 | 42.8 ± 15.7 |
| Gender |  |  |  | <0.001 |  |
| Male | 93,268 (50.2) | 85,748 (48.5) | 122,116 (51.3) |  | 301,132 (50.2) |
| Comorbidities<sup>c</sup> |  |  |  |  |  |
| Essential hypertension | 44,684 (24.1) | 40,164 (22.7) | 51,917 (21.8) | <0.001 | 136,765 (22.8) |
| Chronic cystitis | 533 (0.3) | 669 (0.4) | 740 (0.3) | <0.001 | 1,942 (0.3) |
| Smoking-related diagnoses | 11,003 (5.9) | 12,311 (7.0) | 14,741 (6.2) | <0.001 | 38,055 (6.3) |
| Alcohol use disorders | 4,271 (2.3) | 3,428 (1.9) | 4,991 (2.1) | <0.001 | 12,690 (2.1) |
| Morbid obesity | 1,673 (0.9) | 1,332 (0.8) | 2,402 (1.0) | <0.001 | 5,407 (0.9) |
| Spinal cord injury | 1,451 (0.8) | 1,407 (0.8) | 1,242 (0.5) | <0.001 | 4,100 (0.7) |
| Diabetes mellitus | 31,447 (16.9) | 31,065 (17.6) | 39,392 (16.5) | <0.001 | 101,904 (17.0) |
| Chronic liver disease | 23,456 (12.6) | 21,790 (12.3) | 29,948 (12.6) | 0.016 | 75,194 (12.5) |
| Gout | 20,695 (11.1) | 18,857 (10.7) | 25,572 (10.7) | <0.001 | 65,124 (10.9) |
| Chronic kidney disease | 6,024 (3.2) | 6,062 (3.4) | 6,682 (2.8) | <0.001 | 18,768 (3.1) |
| Pesticide exposures | 310 (0.2) | 253 (0.1) | 231 (0.1) | <0.001 | 794 (0.1) |
| Dyslipidemia | 43,749 (23.6) | 40,130 (22.7) | 56,142 (23.6) | <0.001 | 140,021 (23.3) |

CI, confidence interval; SD, standard deviation; UBC, urinary bladder cancer
<sup>a</sup>The tertile values, in ppm, are as follows: T1 <2.10, T2 ≥2.10 and <2.25, and T3 ≥2.25
<sup>b</sup>The chi-squared test or one-way analysis of variance among tertiles of total hydrocarbons
<sup>c</sup>Comorbidities were before the survival date

**Table 2.** Relative risks of incident UBC by each tertile of nonmethane hydrocarbons exposure and characteristics of the cohorts.

| Characteristics | Tertile of daily cumulative average NMHC<sup>a</sup>, n (%) |  |  | *P*<br>value<sup>b</sup> | Total (N =<br>600,666) |
| --- | --- | --- | --- | --- | --- |
|  | T1 (lowest) (n =<br>187,534) | T2 (n =<br>265,282) | T3 (highest) (n =<br>147,850) |  |  |
| Urinary bladder cancer | 338 | 537 | 671 |  | 1,546 |
| Incident UBC per 100,000 | 180.2 | 202.4 | 453.8 |  | - |
| Relative risk (95% CI) | Reference | 1.12 (0.98–<br>1.29) | 2.52 (2.21–2.87) | <0.001 | - |
| Attributable risk proportion<br>(95% CI) | Reference | - | 40.09% (34.89–<br>45.29%) |  |  |
| Age, years |  |  |  | <0.001 |  |
| 20–29 | 39,585 (21.1) | 64,279 (24.2) | 42,301 (28.6) |  | 146,165 (24.3) |
| 30–39 | 45,246 (24.1) | 65,649 (24.7) | 39,311 (26.6) |  | 150,206 (25.0) |
| 40–49 | 42,090 (22.4) | 57,242 (21.6) | 32,596 (22.0) |  | 131,928 (21.9) |
| 50–59 | 26,150 (13.9) | 31,865 (12.0) | 15,673 (10.6) |  | 73,688 (12.3) |
| 60–69 | 21,549 (11.5) | 24,277 (9.2) | 9,387 (6.3) |  | 55,213 (9.2) |
| ≥70 | 12,914 (6.9) | 21,970 (8.3) | 8,582 (5.8) |  | 43,466 (7.3) |
| Mean ± SD | 44.1 ± 15.5 | 43.2 ± 16.1 | 40.6 ± 15.0 | <0.001 | 42.8 ± 15.7 |
| Gender |  |  |  | <0.001 |  |
| Male | 95,129 (50.7) | 126,984 (47.9) | 79,019 (53.4) |  | 301,132 (50.2) |
| Comorbidities<sup>c</sup> |  |  |  |  |  |
| Essential hypertension | 46,748 (24.9) | 59,929 (22.6) | 30,088 (20.4) | <0.001 | 136,765 (22.8) |
| Chronic cystitis | 861 (0.5) | 737 (0.3) | 344 (0.2) | <0.001 | 1,942 (0.3) |
| Smoking-related diagnoses | 13,678 (7.3) | 16,352 (6.2) | 8,025 (5.4) | <0.001 | 38,055 (6.3) |
| Alcohol use disorders | 4,543 (2.4) | 4,904 (1.8) | 3,243 (2.2) | <0.001 | 12,690 (2.1) |
| Morbid obesity | 1,606 (0.9) | 2,292 (0.9) | 1,509 (1.0) | <0.001 | 5,407 (0.9) |
| Spinal cord injury | 1,606 (0.9) | 1,739 (0.7) | 755 (0.5) | <0.001 | 4,100 (0.7) |
| Diabetes mellitus | 34,893 (18.6) | 44,803 (16.9) | 22,208 (15.0) | <0.001 | 101,904 (17.0) |
| Chronic liver disease | 25,584 (13.6) | 32,325 (12.2) | 17,285 (11.7) | <0.001 | 75,194 (12.5) |
| Gout | 22,628 (12.1) | 27,188 (10.2) | 15,308 (10.4) | 0.001 | 65,124 (10.9) |
| Chronic kidney disease | 6,231 (3.3) | 8,850 (3.3) | 3,687 (2.5) | <0.001 | 18,768 (3.1) |
| Pesticide exposures | 418 (0.2) | 273 (0.1) | 103 (0.1) | <0.001 | 794 (0.1) |
| Dyslipidemia | 46,802 (25.0) | 61,122 (23.0) | 32,097 (21.7) | 0.001 | 140,021 (23.3) |

CI, confidence interval; NMHC, nonmethane hydrocarbon; SD, standard deviation; UBC, urinary bladder cancer
<sup>a</sup>The tertile values, in ppm, are as follows: T1: <0.26, T2: ≥0.26 and <0.34, and T3: ≥0.34
<sup>b</sup>The Chi-squared test or one-way analysis of variance among tertiles of nonmethane hydrocarbons
<sup>c</sup>Comorbidities were before the survival date

**Figure 1.**
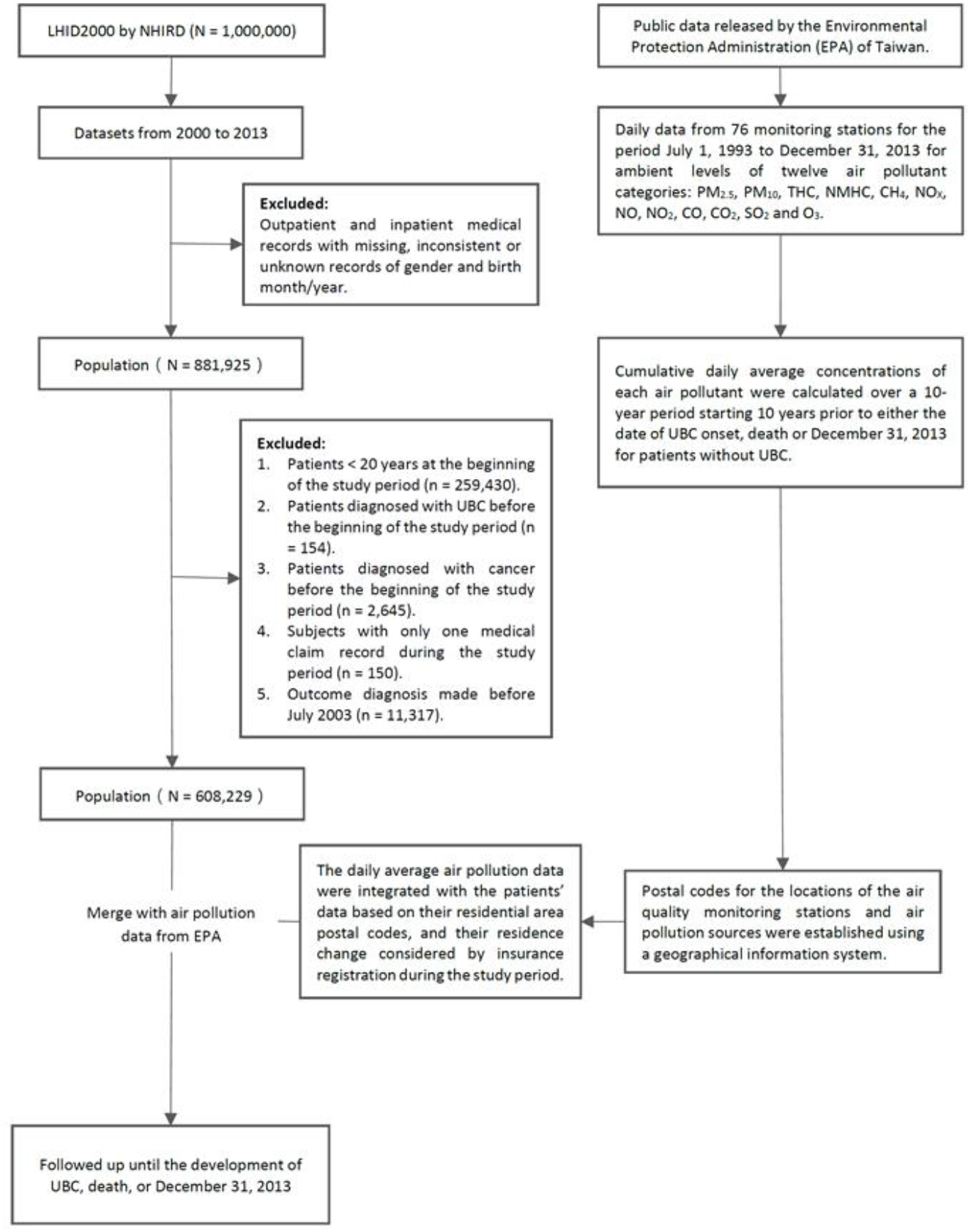
The study flow diagram.

### Air pollution exposure

In the current study over the 10-year exposure period, the mean daily average of THC concentration was 2.24 ppm (SD = 0.14); NMHC was 0.29 ppm (SD = 0.09). Table S1 shows the Pearson’s correlation analysis for the 12 air pollutants over a 10-year exposure period. The absolute value of the correlation coefficients <0.3 denote a low correlation strength, which qualifies as the controlling pollutants in the multiple-pollutant models of the targeted pollutants, THC and NMHC. THC with SO_2_ (r = 0.089), O_3_ (r = -0.296), PM_10_ (r = -0.223), and PM_2.5_ (r = -0.279), while NMHC with SO_2_ (r = 0.156) and CH_4_ (r = 0.142) (Supplementary Table S1). The summary statistics of the air pollutants over a 10-year exposure period are shown in Supplementary Table S2. The distributions of daily average concentrations of air pollutants over the 10-year exposure period are shown in Supplementary Fig. S1 and S2 (SO_2_, CO_2_, CO, O_3_, PM_10,_ and PM_2.5_, are shown in Fig. S1; NOx, NO, NO_2_, THC, NMHC, and CH_4_ in Fig. S2).

### Ambient air THC, NMHC exposure, and incident UBC

Among the target pollutant concentrations to which all cohort participants were exposed, pollutant levels were categorized into tertiles, with T1 being the lowest and T3 being the highest. At the end of the follow-up period, prolonged exposure to THC increased the number of newly diagnosed UBC in a dose-dependent manner: 60.3 cases per 100,000 individuals under T1 concentration exposure, 203.7% under T2 exposure, and 450.8 under T3 exposure (Table 1). For T2 exposure, the relative risk (RR) was 3.38 (95% CI, 2.73–4.17) when compared to that of T1 exposure. Moreover, when comparing T3 to T1, the RR was 7.48 (95% CI, 6.16–9.08) (Table 1). Ambient air NMHC exposure was associated with 180.2/100,000 over the entire study period for the lowest T1 exposure; 202.4 for T2; and 453.8 for T3, indicating a dose-dependent effect (Table 2). While the RR between T1 and T2 did not reach statistical significance, the RR was 2.52 (95% CI, 2.21–2.87) for T3 compared to T1 (Table 2).

Table 3, Table 4, and Supplementary Table S3 present the single- and multiple-pollutant models for per 0.14 ppm or 0.09 ppm increase in THC or NMHC on different stratifications. We fitted two-pollutant models for THC by controlling for the concomitant exposure to SO_2_, O_3_, PM_10_, or PM_2.5_, and the three-pollutant model for THC while controlling for SO_2_ and O_3_. Two-pollutant models for NMHC were used to control for the concomitant exposure to SO_2_ or CH_4,_ and the three-pollutant model for NMHC while controlling for SO_2_ and CH_4_.

**Table 3.** **Adjusted hazard ratios of developing urinary bladder cancer during long-term THC or NMHC air pollutants exposure at a standard deviation (SD) increment controlled for PM_2.5_ and other air pollutants**

| Pollutant category | Controlling pollutant<sup>a</sup> | Adjusted HR<sup>b</sup> (95% CI) |
| --- | --- | --- |
| THC (0.14 ppm increase) | - | 1.77 (1.69–1.87)<sup>‡</sup> |
|  | SO<sub>2</sub> | 1.75 (1.67–1.85)<sup>‡</sup> |
|  | O<sub>3</sub> | 1.61 (1.54–1.69)<sup>‡</sup> |
|  | PM<sub>10</sub> | 2.01 (1.90–2.12)<sup>‡</sup> |
|  | PM<sub>2.5</sub> | 2.10 (1.99–2.22)<sup>‡</sup> |
|  | SO<sub>2</sub>, O<sub>3</sub> | 1.59 (1.52–1.68)<sup>‡</sup> |
| NMHC (0.09 ppm increase) | - | 1.38 (1.31–1.44)<sup>‡</sup> |
|  | SO<sub>2</sub> | 1.36 (1.30–1.43)<sup>‡</sup> |
|  | CH<sub>4</sub> | 1.19 (1.14–1.25)<sup>‡</sup> |
|  | SO<sub>2</sub>, CH<sub>4</sub> | 1.16 (1.11–1.21)<sup>‡</sup> |

HR, hazard ratio; CI, confidence interval; SO<sub>2</sub>, sulfur dioxide; O<sub>3</sub>, ozone; PM<sub>10</sub>, particulate matter <10 µm in size; PM<sub>2.5</sub>, particulate matter <2.5 µm in size; CH<sub>4</sub>, methane
<sup>a</sup>Additional pollutants were added into the pollutant models for multiple analysis only when Pearson's correlation coefficient was <0.3
<sup>b</sup>Cox regression models were adjusted for age, gender, essential hypertension, chronic cystitis, smoking-related diagnoses, alcohol use disorders, morbid obesity, spinal cord injury, chronic liver disease, diabetes mellitus, gout, chronic kidney disease, pesticide exposures, dyslipidemia, lag 0–2, season, and ambient temperature, controlled pollutants (weak correlation with THC or NMHC)
<sup>‡</sup>p <0.001

**Table 4.**
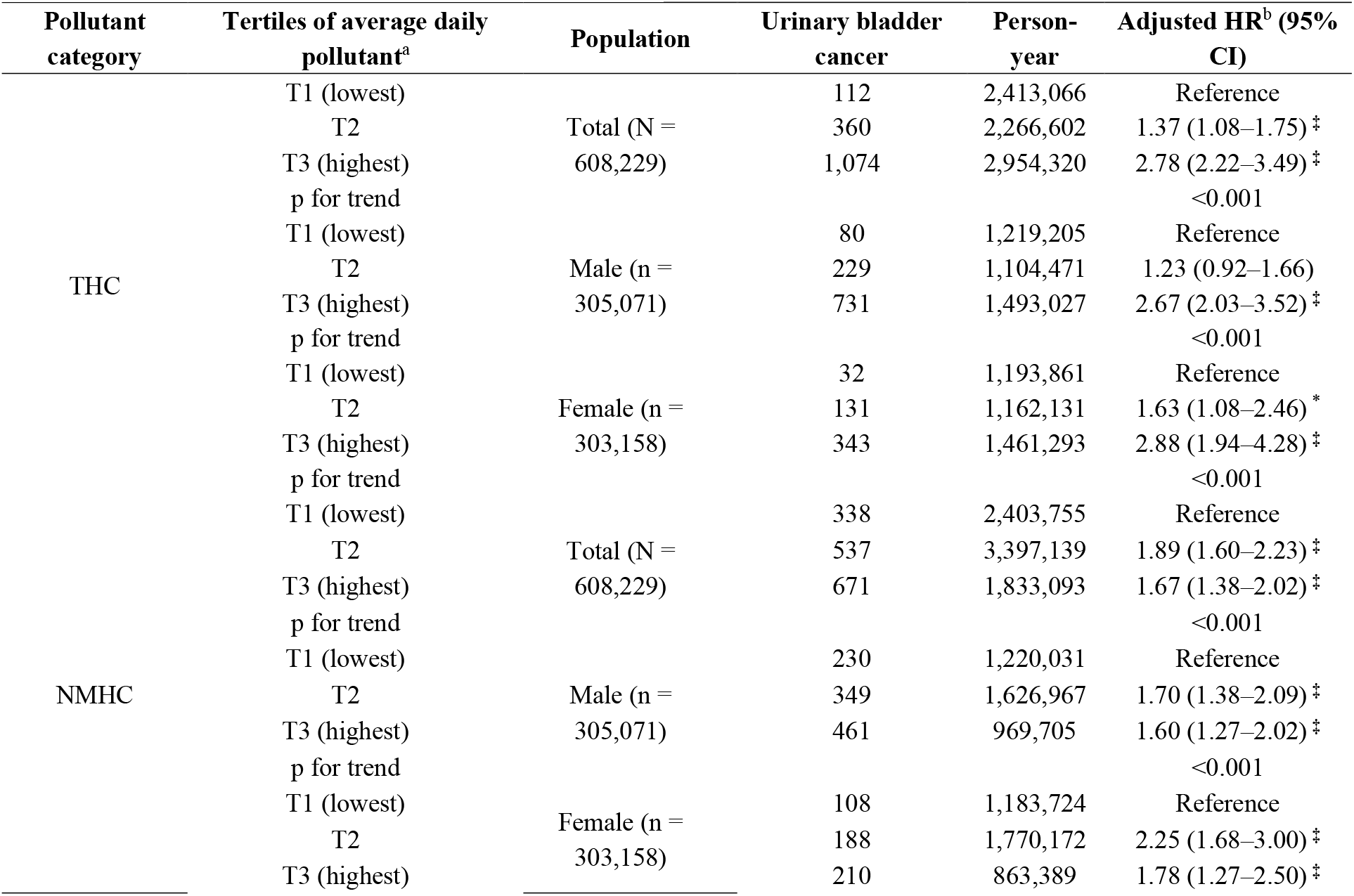
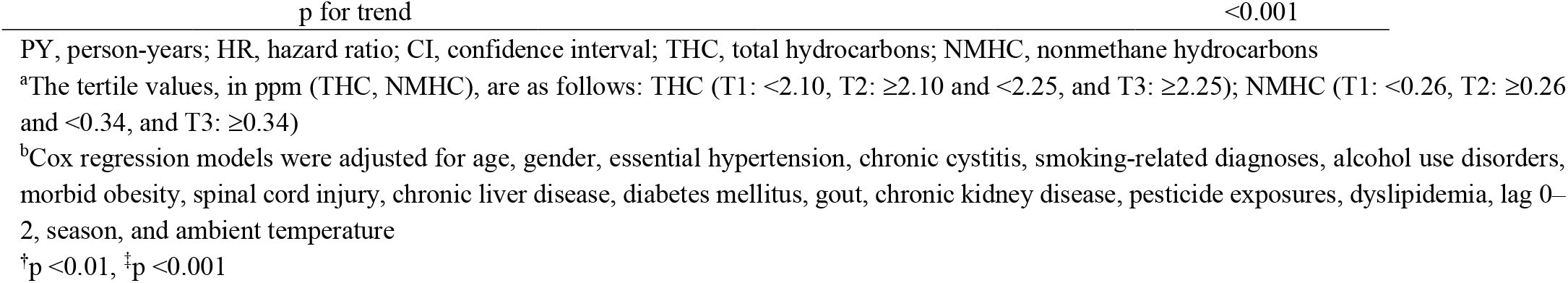
**Cox regression-derived adjusted hazard ratios for incident urinary bladder cancer associated with each tertile of ambient THC or NMHC exposure stratified by gender**

Without controlling for confounding air pollutants, the adjusted hazard ratio (HR) for UBC development was 1.77 (95% CI, 1.69–1.87; *p* <0.001) per 0.14 ppm increase of THC; after controlling for PM_2.5_, adjusted HR was even higher at 2.10 (95% CI, 1.99–2.22). The adjusted HR was 1.38 (95% CI, 1.31–1.44; *p* <0.001) per 0.09 ppm increase in ambient NMHC concentration. After controlling for SO_2_ and CH_4_, the adjusted HR was 1.16 (95% CI, 1.11– 1.21). Ambient NMHC controlling for the most important confounder of air pollutant SO_2_ resulted in an adjusted HR of 1.36 (1.30–1.43, *p* <0.001) (Table 3).

Table 4 presents the Cox proportional hazards regression analysis of the two targeted pollutant categories divided into three tertiles. The lowest tertile was used as the reference in each case, and the estimated HRs were adjusted for age, sex, lag 0–2, season, ambient temperature, and comorbidities. These results were consistent with those obtained from earlier multivariate analyses. Our research shows that the overall population exposure to the highest tertile (T3) of THC significantly increased the risk of UBC, with an adjusted HR (95% CI) of 2.78 (2.22–3.49; p <0.001) (Table 4). Moreover, for those exposed to average daily levels of T3, the risk of UBC was higher than that exposed to T1. For THC, the adjusted HRs of UBC for T3 were 2.67 (2.03–3.52; p <0.001) for males and 2.88 (1.94–4.2; p <0.001) for females. When data on sex were stratified or merged for analysis, statistically significant correlations of adjusted HRs were measured for T2 and T3 compared with T1. The analysis demonstrated an association between the targeted pollutants and UBC risk.

### Sensitivity analyses

To assess the gender-specific differences and the confounding effect of diabetes mellitus status on the association between the targeted pollutants and UBC cancer development, and to reveal any unexpected hidden relationships, we performed sensitivity analyses to compute causal effects only within the underlying strata. The results demonstrated no unexpected relationship, sex-specific differences, or ameliorating impact in non-diabetics (Supplementary Tables S3 and S4).

Before controlling for other pollutants, newly diagnosed UBC in males was significantly positively associated with the daily average concentration over the 10-year period for THC and NMHC with adjusted HR (95% CI) of 1.72 (1.62–1.83; *p* <0.001) and 1.33 (1.26–1.41; *p* <0.001) (Supplementary Table S3). Moreover, in females, the newly diagnosed UBC for THC and NMHC with adjusted HR (95% CI) was 1.88 (1.72–2.05; *p* <0.001) and 1.47 (1.35–1.59; *p* <0.001). Among them, THC controlling for PM_2.5_ resulted in adjusted HRs for males and females of 2.07 (1.94–2.21; *p* <0.001) and 2.26 (2.06,2.49; *p* <0.001), respectively. NMHC controlling for SO_2_ resulted in adjusted HRs for males and females of 1.31 (1.24–1.39; *p* <0.001) and 1.47 (1.36–1.59; *p* <0.001), respectively.

Before controlling for other pollutants, newly diagnosed UBC in people with diabetes mellitus was significantly positively associated with the daily average concentration over the 10-year period for THC and NMHC, with adjusted HRs (95% CI of 2.03 [1.82–2.26; *p* <0.001]) and 1.51 (1.37–1.67; *p* <0.001), respectively (Supplementary Table S4). Moreover, the newly diagnosed UBC without diabetes mellitus for THC and NMHC with adjusted HR was 1.71 (1.61–1.81; *p* <0.001) and 1.34 (1.27–1.41; *p* <0.001), respectively. Among them, THC controlling for PM_2.5_ resulted in adjusted HRs for people with and without diabetes mellitus of 2.55 (2.26–2.87; *p* <0.001) and 2.02 (1.90–2.14; *p* <0.001), respectively. NMHC controlling for SO_2_ resulted in adjusted HRs for people with and without diabetes mellitus of 1.49 (1.34– 1.65]; *p* <0.001) and 1.33 (1.26–1.40; *p* <0.001), respectively.

### Cumulative incidences of UBC compared between different tertiles

In our findings, we observed slight changes in the effects of THC and NMHC after controlling for other pollutants, and the directions of the effect estimates did not change, suggesting that our findings were robust against potential confounders. The appropriateness of the Cox proportional hazards model is supported by the plot in the upper panel of Fig. 2, showing the log (-log [survival function]) versus the log of survival time. Cumulative UBC incidence for the targeted pollutants was assessed using the Kaplan–Meier method (Fig. 2, lower panel), presenting a clear trend of increased UBC risk with increased exposure to each targeted pollutant. Over the entire follow-up period, the incidence of UBC cases per 100,000 people by T1/T2/T3 exposure to THC was 60.3, 203.7, and 450.8, respectively; it was 180.2/202.4/453.8 per 100,000, corresponding to NMHC T1/T2/T3 exposure, respectively. In addition, statistically significant differences in UBC occurrence were observed among the tertiles of the targeted pollutant categories (log-rank test; *p* <0.001). A visualization summary to display the dose-response effect by escalated exposure in tertile is displayed in Figure 3.

**Figure 2.**
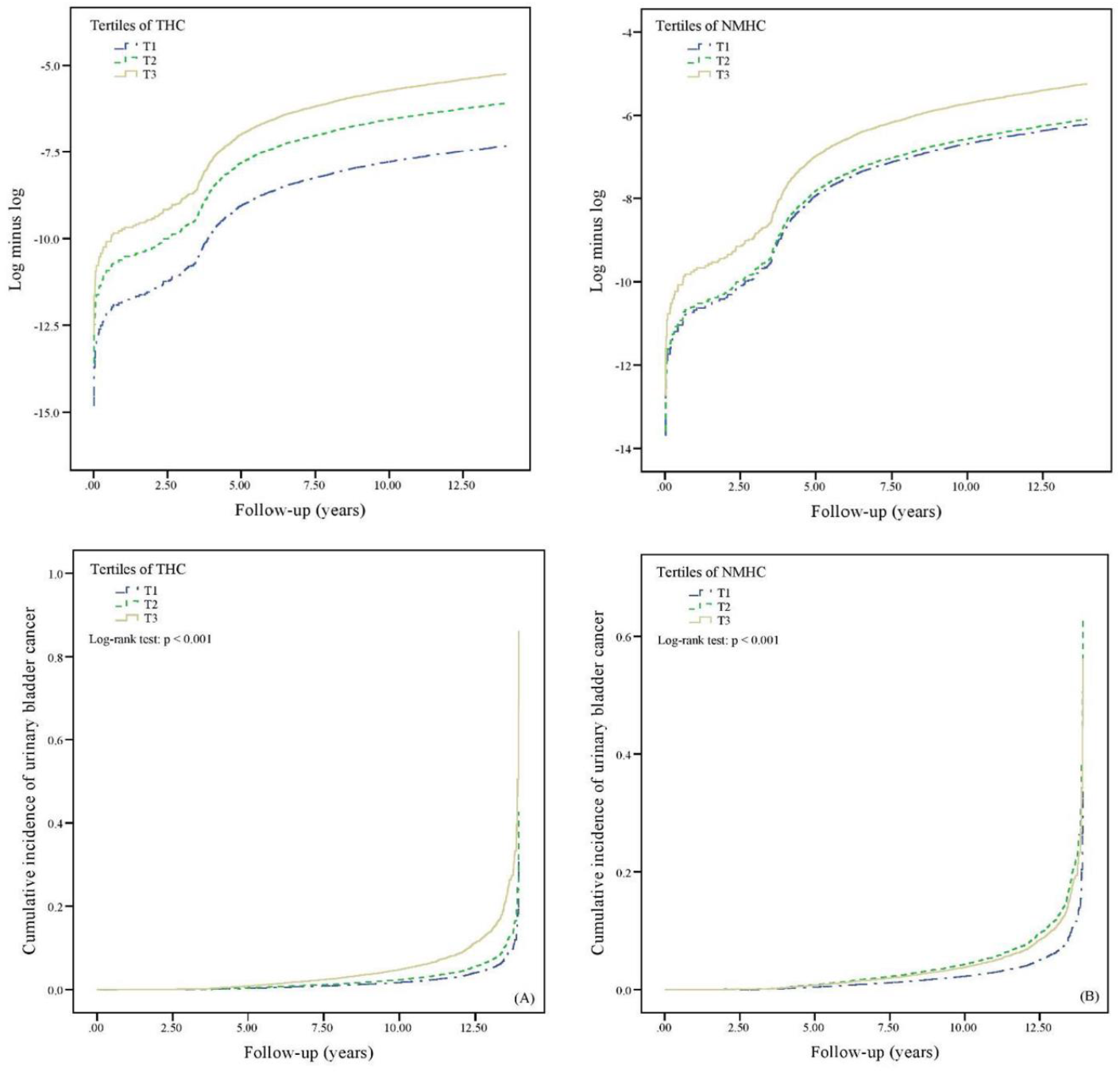
The plot of log (-log [survival function]) versus log of survival time by ambient air THC and NMHC pollutants, and the cumulative incidence of urinary bladder cancer in individuals among THC and NMHC pollutants’ tertile. Upper panel: To evaluate the proportional hazards (PH) assumption involving the comparison of estimated -ln (-ln) survival curves over different THC and NMHC tertiles, the plot of log (-log [survival function]) versus log of survival time in THC and NMHC air pollutants was constructed. The graphical approach showing parallel curves over time provides support of the PH assumption. Lower panel: Cumulative incidence of urinary bladder cancer for individuals among tertiles of THC and NMHC pollutants. The tertile values, in ppm (THC, NMHC), are as follows: THC (lowest tertile, T1: <2.10; medium tertile, T2: ≥2.10 and <2.25; and highest tertile, T3: ≥2.25); NMHC (lowest tertile, T1: <0.26; medium tertile, T2: ≥0.26 and <0.34; and highest tertile, T3: ≥0.34).

**Figure 3.**
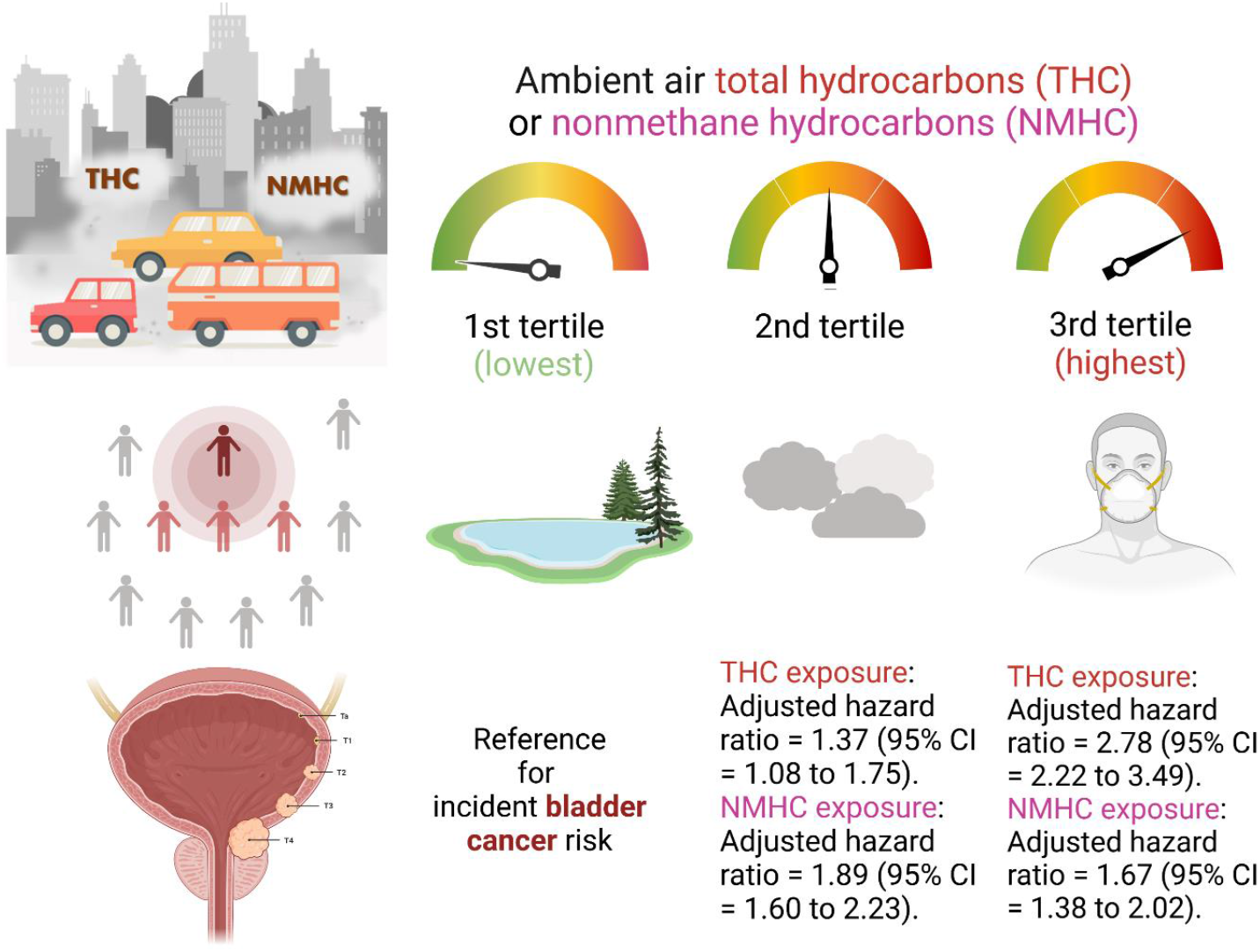
**A schematic diagram summarizing the study results of the risk of developing urinary bladder cancer comparing three tertiles of ambient air concentration of either NMHC or THC. The magnitudes of risk show a significant exposure-response relationship**.

## Discussion

This nationwide population-based cohort study linked national insurance claims data to open government data to investigate the association between long-term exposure to ambient air pollutants, THC or NMHC in Taiwan, and UBC risk. Our novel analysis shows a positive correlation between exposure to hydrocarbons (THC or NMHC) in the ambient air for ten years and UBC risk in people aged ≥20 years. Each additional unit of SD (0.14 ppm; 0.09 ppm) concentration of THC and NMHC increases the risk of bladder cancer by 77% and 38%, respectively. Furthermore, these relationships did not change after the sensitivity analysis of sex or the presence or absence of diabetes. A large body of epidemiological evidence has indicated that diabetes is an independent risk factor for increased rates of heterogeneous types of cancer occurrence and death. The incidence and mortality of various types of cancer, including UBC, have a modest increase in patients with diabetes ^25^. The magnitude of increase of the risk of UBC mortality in people with diabetes in terms of the HR, after being adjusted for baseline age, smoking status, and body mass index, was 1.40 (95% CI, 1.01–1.96). Therefore, we performed sensitivity analyses, and observed a consistent effect on patients without diabetes: the association in our study remained after controlling for simultaneous exposure to other pollutants, particularly PM_2.5_.

Many studies have demonstrated that air pollution adversely impacts health, and may result in problems including cancer; however, few have focused on specific pollutants in ambient air ^2,6,16,21,23,26-28^. Most studies investigated the role of PM_2.5_, NO_2_, NOx, SO_2_, and respirable elemental carbon as a proxy for diesel exhaust in the overall incidence of cancers, including UBC ^2,16,23^. By contrast, our study presents a novel analysis of the association between the long-term exposure to ambient NMHC and THC and the risk of UBC. VOCs, including benzene and formaldehyde, in diesel engine exhaust emissions can be positively correlated with THC emissions, contributing to aggravated ground-level O_3_ pollution when intense solar radiation and high temperatures and low humidity occur. Our prespecified models demonstrated that after controlling for PM_2.5_; health comorbidities; and season and ambient temperatures, each 0.14 ppm increase in THC concentrations would lead to a two-fold increase in the risk of incident UBC in people without diabetes mellitus (adjusted HR, 2.02; 95% CI, 1.90–2.14). When replacing PM_2.5_ with SO_2_ and O_3_, an increase of 54% in the risk of developing UBC was still observed (adjusted HR, 1.54; 95% CI, 1.46–1.63).

A recent systematic review and meta-analysis study pointed out that petroleum industry work was associated with an increased risk of various cancers, including UBC (effect size = 1.25, 95% CI: 1.09–1.43) ^29^. A Spanish case-control study indicated that living more than 40 years in a city with more than 100,000 inhabitants was associated with an increased risk for UBC (OR = 1.30, 95% CI: 1.04–1.63) ^24^. Emissions of PAHs and diesel from industries near the residence, as evaluated by experts, were associated with an increased risk (OR = 1.29, 95% CI: 0.85–1.98) ^24^. In addition, previous research has shown that, for urban bus drivers and tramway employees who were employed for >3 months, the risk of bladder cancer (standardized incidence ratio [SIR] = 1.4, 95% CI: 1.2–1.6) was significantly increased ^7^. The SIRs and 95% CIs for bladder cancer in road transportation workers compared with those in the whole population were 1.26 and 1.03–1.52, respectively ^6^. Nevertheless, assuming occupational exposure, such as among road transportation workers, motor vehicle mechanics and repairers, garage mechanics, underground mines workers, and petroleum industry workers, as a proxy to ambient air pollution may be invalid since it is possible that the route of entry into the human body might have been via skin contact or even through the mucosal organs and not from inhalational hazard sources. Similarly, using petrol station density or annual industrial waste gas emissions to represent exposure to air pollution may also be groundless due to poor representation of specific pollutants. Thus, our study specifically targeted air pollutants such as THC and NMHC to offer new evidence on the impact of these precise components on the development of UBC.

In addition, we used a multiple-pollutant model to control the association between hydrocarbons and the incidence of bladder cancer. We found that, compared with the THC single-pollutant model, THC controlling for PM_2.5_ has a higher risk of bladder cancer, while the risk of THC controlling for O_3_ is lower. Although previous studies used mortality rather than new cancer incidence as endpoints, they still show that PM_2.5_ has a significant deleterious effect on bladder cancer ^18,19^. However, no effective research on O_3_ and bladder cancer has been conducted. In an analysis of 623,048 ACS CPS-II participants in the United States, there was a significant adverse association between PM_2.5_ and bladder cancer mortality (HR per 4.4 µg/m^3^, 1.13; 95% CI, 1.03–1.23; N = 1324) but no association with NO_2_ or O_3_ ^18^. Thus, it can be seen that in the multiple-pollutant model, PM_2.5_ may increase the risk of target pollutants in bladder cancer. However, O_3_ is negatively correlated with other air pollutants, which may be due to the broader spatial pattern of concentration, making it unable to capture fine-scale variation and scavenging effects in urban areas ^18,30^.

Sufficient or convincing evidence about UBC carcinogenesis is associated with genetic susceptibility, cigarette smoking, diet, *Schistosoma haematobium* infestation, and environmental pollution exposure ^31^. However, the biological mechanism of carcinogenesis related to ambient air pollution remains unclear. Air pollution contains various mutagens and carcinogens, which may play a role in chronic systemic inflammation, oxidative stress, and DNA damage in tissues other than the lungs ^15,16,32-34^. Accordingly, exposure to ambient air pollution may induce carcinogenic genotoxic effects in the urinary bladder, providing biological plausibility of an association between hydrocarbon and UBC. Multiple lines of indirect evidence have shed light on the mechanistic explanation for ambient air THC and NMHC-related UBC carcinogenesis. The toxic potential of environmental pollutants can induce oxidative stress, inflammatory potential, DNA damage, and aryl hydrocarbon receptor (AhR) agonist activity ^33-36^. AhR is an important target for environmental carcinogens. Recent basic science research has focused on the molecular mechanisms of the environmental xenobiotics-activated AhR/cytochrome P450 pathway ^36-39^. An elegant study demonstrated that AhR activity was primarily activated by open burning, vehicle exhaust, and naphthalene-derived secondary organic aerosols ^40^. Moreover, in cell line experiments, AhR activation was associated with tumor grade, cancer stage, and progression of UBC ^41^. Hence, before we have more convincing and direct evidence for the ambient air THC and NMHC-induced carcinogenesis of UBC, the AhR/CYP1A1 pathway activation, in addition to the canonical oxidative, inflammatory, and DNA-damage mechanisms, should be investigated in the THC and NMHC carcinogenesis models in future research.

The strengths of this study are as follows. First, this is a nationwide study using a large population derived from the NHIRD, which contains medical care data for 2,296 million people (99% of Taiwan’s population) under the National Health Insurance Plan. Second, this study is based on a 10-year long-term follow-up, which can provide adequate follow-up time to assess UBC development. Third, few epidemiological studies have assessed the association between airborne hydrocarbons and UBC in Asia. Moreover, many studies have dealt with an increase in mortality rather than cancer incidence. These endpoints are different in that the increase in mortality from UBC is due to the promoting effect of pollutants on cancer progression, whereas an increase in UBC incidence indicates the presence of the carcinogenic effect of air pollutants. Fourth, we considered and included the most important known risk factors for UBC in our calculation models and examined the possible synergistic effects between air pollutants. Finally, this study explored the association between hydrocarbons and UBC through sex and diabetes stratification.

This linkage dataset cohort study has several limitations. First, because study subjects might relocate to a new place, exposure to air pollutants may change accordingly if the geographic location has a different level of pollutant concentration. The impact of this transition was difficult to measure. However, for most subjects in the cohort, a fixed postal address for one exposure time was reliable. Second, we could not measure occupational exposure to other hazardous chemicals in the workplace. Lastly, we also lack individual dietary habit records to measure the possible exposure to food chain carcinogens in the current study. However, we believe that the magnitude of the impact of these limitations would not be large enough to alter the direction of the risk of developing UBC in this large population-based cohort study.

## Conclusions

This ambispective cohort study offered new evidence that long-term exposure to THC and NMHC may be a risk factor for UBC. The results indicate a possible link between hydrocarbons and UBC risk. Further, in a stratified analysis of the population in Taiwan by gender or diabetes, long-term exposure to the two target pollutant categories is associated with an increased risk of UBC. Specifically, the leading cause and mechanism of the disease remain unclear. Therefore, the prevention method is to avoid dangerous factors as much as possible or reduce exposure to hazardous environments. Early detection and treatment are also key points for prevention and treatment. With the global aging trend, the prevalence and burden of UBC may also increase. Acknowledging the pollutant sources harmful to health can provide information for risk management strategies and help decision-makers formulate more targeted air pollution regulations. These findings may have important public health implications for preventing UBC.

## Methods

### Ethics approval and consent to participate

This study was approved after a full ethical review by the Institutional Review Board (IRB) of the China Medical University, Taichung, Taiwan (approval number: CMUH104-REC2-115 [CR-6]). In addition, because de-identified/anonymized data were used from the NHIRD, the IRB waived the requirement to obtain informed consent from the study participants. All experiments were performed according to confidentiality guidelines set forth by the Taiwan Personal Information Protection Act regulations. The entire study was conducted in accordance with the Declaration of Helsinki.

### Data sources for linkage dataset ambispective cohort research

Health data were obtained from the Longitudinal Health Insurance Database 2000 (LHID2000) within the NHIRD, including claims data for 1 million randomly selected individuals, from 1996 to 2013 ^42^. The NHIRD, established in 1996 in Taiwan, contains healthcare data of 22.96 million people (99% of Taiwan’s population) under a universal health insurance program, including all claims data (ambulatory care claims and inpatient claims) and prescriptions dispensed at pharmacies, the registry for beneficiaries, registry for medical facilities, and registry for medical specialists. To establish demographic characteristics for research, patient-level information is gathered by linking these data files using the identification number of insured individuals. As recorded in the database, each individual’s health and disease status was assigned an International Classification of Disease, Ninth Revision, Clinical Modification (ICD-9-CM) until recently, when ICD-10-CM was implemented. To enhance the reliability of the NHIRD data, the observation period was set as 2000–2013.

In addition, the Environment Resource Datasets^43^ are publicly available from open government data. This dataset was obtained by the Environmental Protection Administration of Taiwan, which determined ambient pollutants and temperatures at 76 monitoring stations across Taiwan, from 1993 to 2013. In this linkage database research, we used the postal code location as a proxy for the residence location from the NHIRD dataset and matched the postal code locations to the corresponding air quality monitoring stations in the Environmental Protection Administration (EPA) Open Dataset.

### Study design and study population

A nationwide linkage database ambispective cohort design was used for this study from January 1, 2000, to December 31, 2013. The selection of the study subjects is depicted in Fig. 1. Among the 1 million subjects in the LHID2000 database, individuals aged 20 years and above were enrolled on January 1, 2000 (n = 622,495). Those with missing or unknown records for sex and birth were excluded. Those with UBC (n = 154) and cancer from any other primary site (n = 2,645) diagnosed before the beginning of the study period; those with only one claim record during the study period (n = 150); and to avoid the reverse causation bias, those with outcome diagnosis made before July 2003 (n = 11,317) were excluded. Ultimately, 608,229 subjects were included in the study.

### Exposure modeling

We established from our research hypothesis to measure exposure to the targeted pollutants. Our devised exposure model for this study incorporated ambient concentrations of targeted pollutants over time while simultaneously addressing personal exposures tracked with the residential information and the duration of contact as input variables to estimate the cumulative individual exposure from inhalation. We have previously reported a similar exposure modeling approach which has drawn acceptance from the exposure and outcomes research community ^44^. We determined the concentrations of 12 ambient air pollutants monitored by the EPA in Taiwan over a pre-specified study period. The study targets were THCs and NMHCs. To examine the association between long-term exposure to targeted air pollutants and the development of newly diagnosed UBC, we measured the risk magnitude after controlling for other non-targeted pollutants over the exposure period. Non-targeted pollutants were included in the subsequent multiple-pollutant analyses. These were selected based on weak correlations (Pearson’s correlation coefficients <0.3) of target pollutants with ten other monitored air pollutants: sulfur dioxide (SO_2_); ozone (O_3_); carbon monoxide (CO); carbon dioxide (CO_2_); nitrogen oxides (NO_X_); nitrogen monoxide (NO); nitrogen dioxide (NO_2_); particulate matter <10 μm in size (PM_10_); particulate matter <2.5 μm in size (PM_2.5_); and methane (CH_4_) (Supplementary Table S1). Daily air quality data were collected at 76 monitoring stations from July 1, 1993, to December 31, 2013, and maintained by the EPA ^**43**^. The locations where air pollutants were recorded were selected to form an integrated geographic information system. Using this system, each study patient was linked to the appropriate monitoring region by postal code, and the change in residence was considered through insurance registration during the study period. A patient’s long-term exposure to each air pollutant was defined as the cumulative concentration during the measurement period (i.e., ten years before the survival date) averaged per day. Therefore, the long-term exposure to each air pollutant (LEAP_ij_) (i = SO_2_, O_3_, CO, CO_2_, NO_X_, NO, NO_2_, PM_10_, PM_2.5_, THC, NMHC, and CH_4_) for a patient living in the region served by the air quality monitoring station j was calculated as follows ^**44**,**45**^:

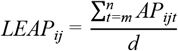

where *APi* is the ambient air pollution level for pollutant category i, *m* is the start date of the measurement period (10 years before the survival date), *n* is the end date of the measurement period (survival date), and *d* is the number of days in the measurement period.

### Study outcomes

From the included population, we identified people who received a first-time diagnosis of either invasive or *in situ* UBC during the study period, based on ICD-9-CM codes 188 for invasive carcinoma and 233.7 for urinary bladder carcinoma-*in-situ*, respectively. Individuals were considered to have UBC if they visited an outpatient clinic ≥3 times with a UBC diagnosis or had been hospitalized because of UBC ^46^. The earliest hospitalization or outpatient visit with UBC diagnosis was assigned as the diagnosis date and served as the newly diagnosed date of UBC for all subsequent analyses. We defined survival (the expected duration of time until the outcome event) with an endpoint date of either UBC diagnosis, death, or December 31, 2013, the final observation date, whichever occurred first.

### Comorbidities as confounding factors for UBC outcome were collected

Information on comorbid conditions of patients was determined from the LHID2000 based on ICD-9-CM codes. Comorbidities considered were essential hypertension (401–405); chronic cystitis (595.1, 595.2); smoking-related diagnosis (305.1, 491.0, 491.2, 492.8, 496, 523.6, 989.84, V15.82, 649.0); alcohol use disorders (265.2, 291, 303, 305.0, 357.5, 425.5, 535.3, 571.0, 571.1, 571.2, 571.3, 980.0, V11.3); morbid obesity (278, 646.1, 649.1, 649.2, V45.86, V65.3, V77.8); spinal cord injury (806, 952, 336.1) ^47^; chronic liver disease (571, 572.2–572.9); diabetes mellitus (249, 250, 648.8, 648.0); gout (274); chronic kidney disease (403, 404, 582.9, 585, 646.2, 792.5, 996.1, 999); pesticide exposures (989.1, 989.2, 989.3, 989.4); and dyslipidemia (272). These were identified and defined according to the diagnostic history collected from at least three outpatient visits or a single hospital admission before the survival date.

### Statistical analysis

The chi-squared test (for categorical variables) and one-way analysis of variance (for continuous variables) were used to test for differences in demographic characteristics and distribution of comorbidities among tertiles of the targeted pollutant concentrations. UBC risk associated with each targeted pollutant category, expressed as hazard ratios (HRs) with 95% confidence intervals (CIs), was examined using Cox proportional hazards regression, considering potential confounders. To control the confounding effects of other pollutants, the possible link between air pollutants was used to assess the effects of multiple pollutants, by controlling others that were based on the selection of weak correlations with other air pollutants (i.e., the absolute value of the correlation coefficients between each of the two air pollutants was lower than 0.3; Supplementary Table S1). To avoid potential collinearity problems, we did not include pollutants with high correlations in the same regression model. The effect of each targeted pollutant on the risk of newly diagnosed UBC was estimated as the adjusted HR for the change in standard deviation (SD) over the follow-up period.

Local research has identified a V/U-shaped relationship between air pollutants and ambient temperature, showing significant effects at both ends of extreme temperatures in the region ^48^. Therefore, to control the impact of weather conditions on air pollution and UBC, the ambient temperature should be one of the confounding factors in the pollutant models. Additionally, to control for short-term pollutant exposure effects, we used a lag of 0–2 days (average concentration levels on the same day of the UBC diagnosis, and one and two days before) for all air pollutants as one of the adjusting factors. Because air pollutant levels vary depending on the weather conditions, adjustment for the season is usually considered an important modifier in ambient air pollution-related biological effects in East Asia ^20^. In the present study, multiple-pollutant models for two targeted pollutants were fitted, the independent effects of each targeted pollutant were adjusted for age, sex, comorbidities, lag of 0–2 days, season (seasonal trends in UBC onset), and ambient temperature were estimated, and other pollutants that showed weak correlations were controlled. The concentration data of the targeted pollutants were divided into three tertiles, T1, T2, and T3, and adjusted HRs with 95% CIs were re-calculated.

Sensitivity analyses examined whether the effects of pollutant categories differed between males and females. In addition, studies have pointed out that diabetes is related to a higher risk of UBC ^49^; we decided to use diabetes stratification to explore whether the pollutant category would have a significant impact on the non-diabetic population. Kaplan–Meier analysis was used to determine the cumulative incidence of UBC, and the log-rank test was used to evaluate the difference among tertiles of concentrations of the target pollutants. The analyses were performed using the Meta-Trial Platform and Statistical Product and Service Solutions (SPSS; Version 22). All statistical tests were two-sided; *p* values of 0.05 were considered statistically significant.

## Data Availability

All data produced in the present study are available upon reasonable request to the authors.

## List of abbreviations

AhR: aryl hydrocarbon receptor
CI: confidence interval
EPA: Environmental Protection Administration
HR: hazard ratio
IDC-9-CM: International Classification of Disease, Ninth Revision, Clinical Modification
NHIRD: National Health Insurance Research Database
NMHC: nonmethane hydrocarbon
OR: odds ratio
PAHs: polycyclic aromatic hydrocarbons
PM: particulate matter
RR: relative risk
T: tertile
THC: total hydrocarbon
UBC: urinary bladder cancer
VOCs: volatile organic compounds

## Data availability statement

All data and related metadata underlying reported findings have been deposited in the public data repository: Mendeley Data (https://data.mendeley.com) with a digital object identifier (DOI) as 10.17632/gtpf8t5r9w.1.

## Author contributions statement

HWZ and ZRT conceived the research question and share the first authorship. VCK, JJPT and CYH supervised the research project. VCK, HWZ, ZRT and HCP were responsible for methodology, software and data curation. VCK, HWZ, ZRT, HCP and YHC analyzed and interpreted the study output. VCK, HWZ, and ZRT did the validation. VCK, HWZ, ZRT and HCP wrote the original draft. VCK, HWZ, and HCP worked on the visualization. VCK reviewed and edited the manuscript. All authors read and approved the final manuscript.

## Additional Information

### Competing interests statement

Han-Wei Zhang, Hsiao-Ching Peng reports equipment and statistical analysis were provided by Biomedica Corporation. Han-Wei Zhang reports a relationship with Biomedica Corporation that includes: board membership. Hsiao-Ching Peng reports a relationship with Biomedica Corporation that includes: employment. Both received support for using the product ‘Meta Trial Software’ to conduct the present research. Both declare that they had full access to all the data in this study and take complete responsibility for the integrity of the data and the accuracy of the data analysis. The other authors declare that they have no competing interests.

### Funding

This research was partially supported by the Ministry of Science and Technology through the Center for Precision Medicine Research of Asia University in Taiwan (grant number: MOST 110-2321-B-468-001).

## Acknowledgements

The authors are grateful to the National Health Insurance Administration; Ministry of Health and Welfare, Taiwan; the NHRI, Taiwan; and the Taiwan National Development Council for kindly providing access to the research data for this study. The interpretation and conclusions contained herein do not represent those of the aforementioned institutions. The conception of the research idea, study design, data analysis, data interpretation, and manuscript preparation were solely performed by the authors.

