## Supplementary Fig. 1 for "Long-term ambient hydrocarbon exposure and incidence of urinary bladder cancer"

Supplementary Figures: 2.

Supplementary Tables: 4.

### Legends of Figures and Tables

**Supplementary Figure S1.** Population distribution of the daily average concentrations of air pollutants ($SO_2$, $CO_2$, CO, $O_3$, $PM_{10}$, and $PM_{2.5}$) over a 10-year exposure period.

**Supplementary Figure S2.** Population distribution of the daily average concentrations of air pollutants ($NO_X$, NO, $NO_2$, THC, NMHC, and $CH_4$) over a 10-year exposure period.

**Supplementary Table S1.** Pearson's correlation analysis for air pollutants over a 10-year exposure period. $CO_2$, carbon dioxide; CO, carbon monoxide; $CH_{4,}$ methane; NMHC, nonmethane hydrocarbons; NO, nitrogen monoxide; $NO_{2,}$ nitrogen dioxide; $NO_X$, nitrogen oxides; $O_3$, ozone; $PM_{10}$, particulate matter <10 μm in size; $PM_{2.5,}$ particulate matter <2.5 μm in size; $SO_2$, sulfur dioxide; THC, total hydrocarbons. [†]Correlation significant at the 0.01 level (two-tailed). Correlation coefficient values of <0.3 denote a low strength of correlation, which qualifies as the controlling pollutant in multiple-pollutant models of targeted pollutants.

**Supplementary Table S2.** Mean and distribution of air pollutants over a 10-year exposure period. SD, standard deviation; 5th, 5 percentile; 95th, 95 percentile; Min, minimum; Max, maximum; IQR, interquartile range; $T_1$, 33.33 percentile; $T_2$, 66.66 percentile; ppb, parts per billion; ppm, parts per million; μg/m$^3$, microgram/cubic meter; $CO_2$, carbon dioxide; CO, carbon monoxide; $CH_{4,}$ methane; NMHC, nonmethane hydrocarbons; NO, nitrogen monoxide; $NO_{2,}$ nitrogen dioxide; NOx, nitrogen oxides; $O_3$, ozone; $PM_{10}$, particulate matter <10 μm in size; $PM_{2.5,}$ particulate matter <2.5 μm in size; $SO_2$, sulfur dioxide; THC, total hydrocarbons.

**Supplementary Table S3.** Sensitivity analysis showing adjusted hazard ratios of developing urinary bladder cancer stratified by sex during long-term THC or NMHC exposure at a standard deviation (SD) increment controlled for $PM_{2.5}$, and other air pollutants

**Supplementary Table S4.** Sensitivity analysis showing adjusted hazard ratios of the incidence of urinary bladder cancer stratified by diabetes mellitus status during long-term THC or NMHC exposure at a standard deviation (SD) increment controlled for $PM_{2.5}$, and other air pollutants.

**Supplementary Figure S1.** Population distribution of the daily average concentrations of air pollutants ($SO_2$, $CO_2$, CO, $O_3$, $PM_{10}$, and $PM_{2.5}$) over a 10-year exposure period.

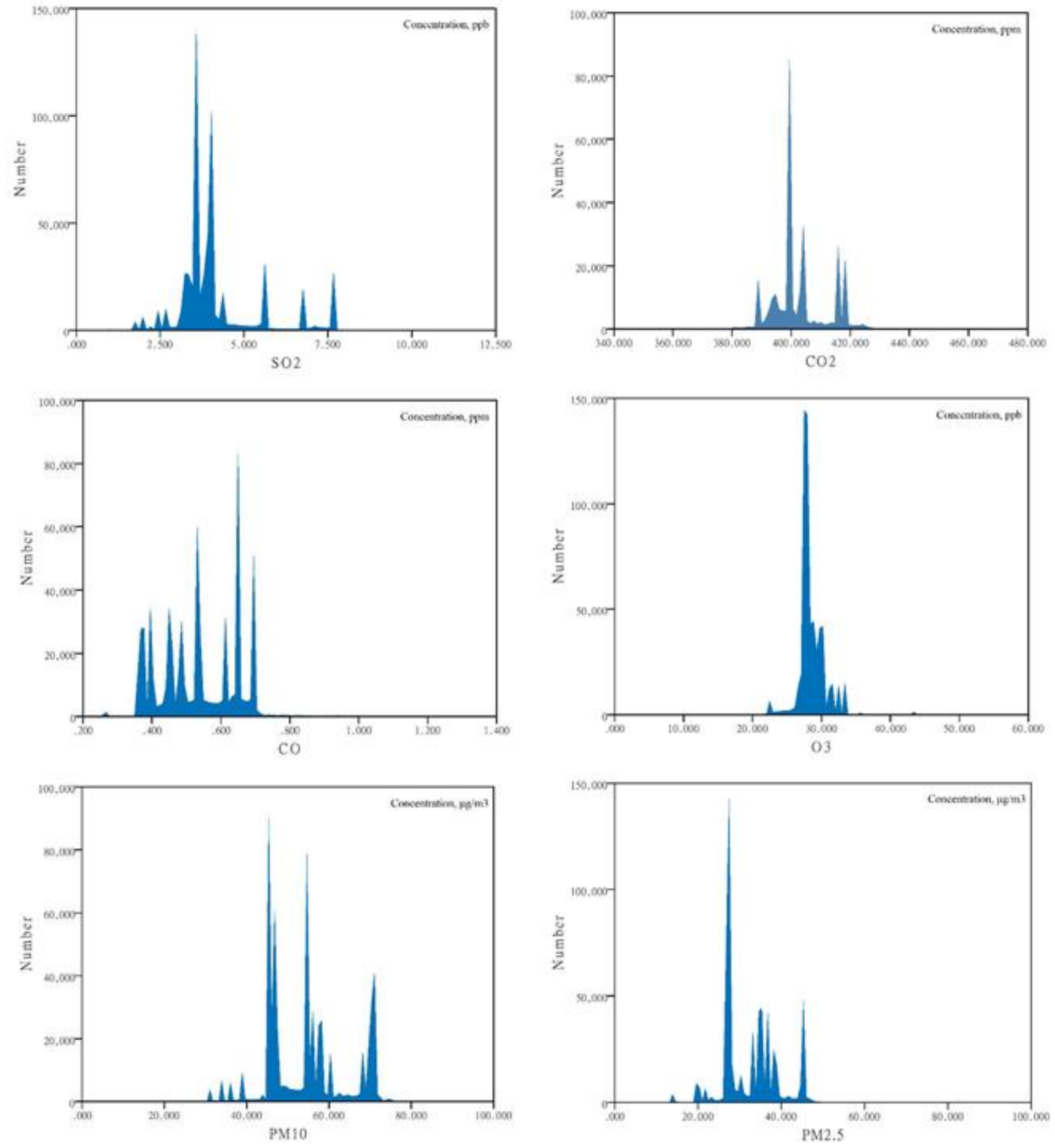

**Supplementary Figure S2.** Population distribution of the daily average concentrations of air pollutants ($NO_X$, NO, $NO_2$, THC, NMHC, and $CH_4$) over a 10-year exposure period.

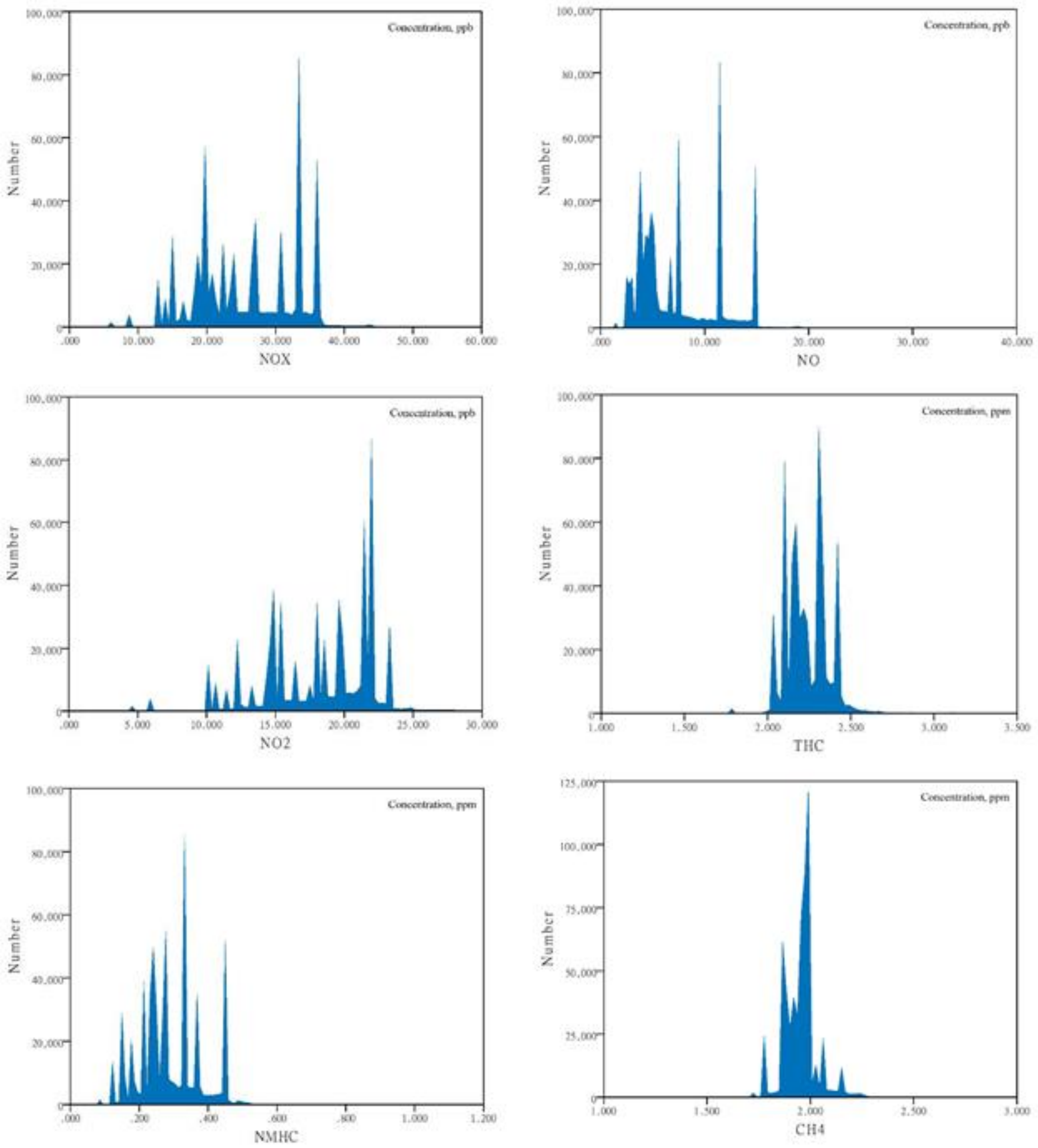

**Supplementary Table S1.** Pearson's correlation analysis for air pollutants over a 10-year exposure period.

| | $SO_2$ | $CO_2$ | CO | $O_3$ | $PM_{10}$ | $PM_{2.5}$ | $NO_X$ | NO | $NO_2$ | THC | NMHC | $CH_4$ |
|---|---|---|---|---|---|---|---|---|---|---|---|---|
| $SO_2$ | 1 | **0.098**† | **0.186**† | **0.059**† | 0.621† | 0.613† | **0.272**† | **0.073**† | 0.449† | **0.089**† | **0.156**† | **-0.050**† |
| $CO_2$ | | 1 | -0.328† | **-0.006**† | 0.527† | 0.413† | **-0.274**† | -0.377† | **-0.154**† | -0.545† | -0.474† | -0.372† |
| CO | | | 1 | -0.602† | -0.319† | **-0.231**† | 0.945† | 0.923† | 0.867† | 0.643† | 0.870† | **0.095**† |
| $O_3$ | | | | 1 | 0.402† | 0.358† | -0.494† | -0.460† | -0.478† | **-0.296**† | -0.481† | **-0.019**† |
| $PM_{10}$ | | | | | 1 | 0.936† | **-0.242**† | -0.427† | **-0.026**† | **-0.223**† | -0.373† | **0.020**† |
| $PM_{2.5}$ | | | | | | 1 | **-0.184**† | -0.387† | **0.045**† | **-0.279**† | -0.349† | **-0.094**† |
| $NO_X$ | | | | | | | 1 | 0.949† | 0.946† | 0.679† | 0.831† | **0.154**† |
| NO | | | | | | | | 1 | 0.795† | 0.752† | 0.879† | **0.231**† |
| $NO_2$ | | | | | | | | | 1 | 0.529† | 0.692† | **0.056**† |
| THC | | | | | | | | | | 1 | 0.750† | 0.743† |
| NMHC | | | | | | | | | | | 1 | **0.142**† |
| $CH_4$ | | | | | | | | | | | | 1 |

$CO_2$, carbon dioxide; CO, carbon monoxide; $CH_4$, methane; NMHC, nonmethane hydrocarbons; NO, nitrogen monoxide; $NO_2$, nitrogen dioxide; $NO_X$, nitrogen oxides; $O_3$, ozone; PM10, particulate matter < 10 μm in size; $PM_{2.5}$, particulate matter < 2.5 μm in size; $SO_2$, sulfur dioxide; THC, total hydrocarbons.

†Correlation significant at the 0.01 level (two-tailed).

Correlation coefficient values of < 0.3 denote a low strength of correlation, which qualifies as the controlling pollutant in multiple-pollutant models of targeted pollutants.

**Supplementary Table S2.** Mean and distribution of air pollutants over a 10-year exposure period.

| | Mean | SD | Median | 5th | 95th | Min | Max | IQR | $T_1$ / $T_2$ cutoff | $T_2$ / $T_3$ cutoff |
|---|---|---|---|---|---|---|---|---|---|---|
| $SO_2$ (ppb) | 4.17 | 1.29 | 3.85 | 2.69 | 7.63 | 12.10 | 0.50 | 0.58 | 3.54 | 4.04 |
| $CO_2$ (ppm) | 402.75 | 9.55 | 398.96 | 388.45 | 418.07 | 463.65 | 343.47 | 8.61 | 398.96 | 404.31 |
| CO (ppm) | 0.54 | 0.12 | 0.53 | 0.37 | 0.69 | 1.32 | 0.25 | 0.20 | 0.47 | 0.61 |
| $O_3$ (ppb) | 28.52 | 2.10 | 27.98 | 26.63 | 32.29 | 52.36 | 1.00 | 1.99 | 27.75 | 28.80 |
| $PM_{10}$ (μg/m$^3$) | 54.47 | 10.02 | 54.54 | 40.02 | 70.75 | 85.20 | 1.00 | 14.03 | 46.72 | 57.50 |
| $PM_{2.5}$ (μg/m$^3$) | 32.44 | 6.81 | 32.32 | 23.02 | 45.49 | 89.43 | 0.64 | 9.38 | 27.45 | 35.13 |
| $NO_X$ (ppb) | 25.78 | 7.52 | 25.98 | 14.11 | 36.22 | 58.53 | 1.00 | 13.86 | 20.59 | 30.74 |
| NO (ppb) | 7.55 | 4.04 | 6.72 | 2.85 | 14.88 | 30.25 | 0.08 | 7.27 | 4.79 | 8.41 |
| $NO_2$ (ppb) | 18.23 | 3.90 | 19.17 | 10.68 | 23.16 | 29.50 | 1.00 | 6.29 | 16.55 | 21.35 |
| THC (ppm) | 2.24 | 0.14 | 2.23 | 2.03 | 2.41 | 3.35 | 1.00 | 0.18 | 2.17 | 2.32 |
| NMHC (ppm) | 0.29 | 0.09 | 0.27 | 0.15 | 0.45 | 1.09 | 0.06 | 0.10 | 0.24 | 0.33 |
| $CH_4$ (ppm) | 1.95 | 0.08 | 1.96 | 1.82 | 2.08 | 2.77 | 1.00 | 0.09 | 1.92 | 1.98 |

SD, standard deviation; 5th, 5 percentile; 95th, 95 percentile; Min, minimum; Max, maximum; IQR, interquartile range; $T_1$, 33.33 percentile; $T_2$, 66.66 percentile; ppb, parts per billion; ppm, parts per million; μg/m$^3$, microgram/cubic meter; $CO_2$, carbon dioxide; CO, carbon monoxide; $CH_4$, methane; NMHC, nonmethane hydrocarbons; NO, nitrogen monoxide; $NO_2$, nitrogen dioxide; NOx, nitrogen oxides; $O_3$, ozone; $PM_{10}$, particulate matter < 10 μm in size; $PM_{2.5}$, particulate matter < 2.5 μm in size; $SO_2$, sulfur dioxide; THC, total hydrocarbons.

**Supplementary Table S3.** Sensitivity analysis showing adjusted hazard ratios of developing urinary bladder cancer stratified by sex during long-term THC or NMHC exposure at a standard deviation (SD) increment controlled for $PM_{2.5}$, and other air pollutants

| **Ambient pollutant category** | **Controlling pollutant**[a] | **Adjusted HR**[b] **(95% CI)** | |
|---|---|---|---|
| | | **Male (n = 305,071)** | **Female (n = 303,158)** |
| THC (0.14 ppm increase) | - | 1.72 (1.62–1.83) ‡ | 1.88 (1.72–2.05) ‡ |
| | $SO_2$ | 1.70 (1.60–1.81) ‡ | 1.87 (1.71–2.04) ‡ |
| | $O_3$ | 1.57 (1.48–1.66) ‡ | 1.72 (1.58–1.87) ‡ |
| | $PM_{10}$ | 1.96 (1.84–2.09) ‡ | 2.10 (1.91–2.30) ‡ |
| | $PM_{2.5}$ | 2.07 (1.94–2.21) ‡ | 2.26 (2.06–2.49) ‡ |
| | $SO_2$, $O_3$ | 1.53 (1.44–1.62) ‡ | 1.75 (1.60–1.91) ‡ |
| NMHC (0.09 ppm increase) | - | 1.33 (1.26–1.41) ‡ | 1.47 (1.35–1.59) ‡ |
| | $SO_2$ | 1.31 (1.24–1.39) ‡ | 1.47 (1.36–1.59) ‡ |
| | $CH_4$ | 1.17 (1.11–1.23) ‡ | 1.25 (1.16–1.35) ‡ |
| | $SO_2$, $CH_4$ | 1.12 (1.06–1.19) ‡ | 1.23 (1.14–1.33) ‡ |

HR, hazard ratio; CI, confidence interval; $SO_2$, sulfur dioxide; $O_3$, ozone; $PM_{10}$, particulate matter < 10 μm in size; $PM_{2.5}$, particulate matter < 2.5 μm in size; $CH_4$, methane.

[a] Additional pollutants were added into the pollutant models for multiple analysis only when |Pearson's correlation coefficient| < 0.3 (Suppl Table S1).

[b] Cox regression models were adjusted for age, gender, essential hypertension, chronic cystitis, smoking-related diagnoses, alcohol use disorders, morbid obesity, spinal cord injury, chronic liver disease, diabetes mellitus, gout, chronic kidney disease, pesticide exposures, dyslipidemia, lag0-2, season, and ambient temperature, controlled pollutants (weak correlation with THC or NMHC).

‡p < 0.001.

**Supplementary Table S4.** Sensitivity analysis showing adjusted hazard ratios of the incidence of urinary bladder cancer stratified by diabetes mellitus status during long-term THC or NMHC exposure at a standard deviation (SD) increment controlled for $PM_{2.5}$, and other air pollutants.

| **Air pollutant category** | **Controlling pollutant**[a] | **Adjusted HR**[b] **(95% CI)** | |
|---|---|---|---|
| | | **Diabetes mellitus (n = 103,317)** | **Non-diabetes mellitus (n = 504,912)** |
| THC (0.14 ppm increase) | - | 2.03 (1.82–2.26) ‡ | 1.71 (1.61–1.81) ‡ |
| | $SO_2$ | 2.00 (1.79–2.23) ‡ | 1.69 (1.60–1.79) ‡ |
| | $O_3$ | 1.82 (1.64–2.02) ‡ | 1.56 (1.48–1.65) ‡ |
| | $PM_{10}$ | 2.34 (2.09–2.64) ‡ | 1.92 (1.81–2.04) ‡ |
| | $PM_{2.5}$ | 2.55 (2.26–2.87) ‡ | 2.02 (1.90–2.14) ‡ |
| | $SO_2$, $O_3$ | 1.80 (1.61–2.01) ‡ | 1.54 (1.46–1.63) ‡ |
| NMHC (0.09 ppm increase) | - | 1.51 (1.37–1.67) ‡ | 1.34 (1.27–1.41) ‡ |
| | $SO_2$ | 1.49 (1.34–1.65) ‡ | 1.33 (1.26–1.40) ‡ |
| | $CH_4$ | 1.31 (1.19–1.44) ‡ | 1.16 (1.10–1.21) ‡ |
| | $SO_2$, $CH_4$ | 1.26 (1.14–1.39) ‡ | 1.12 (1.07–1.18) ‡ |

HR, hazard ratio; CI, confidence interval; $SO_2$, sulfur dioxide; $O_3$, ozone; $PM_{10}$, particulate matter < 10 μm in size; $PM_{2.5}$, particulate matter < 2.5 μm in size; $CH_4$, methane.

[a]Additional pollutants were added into the pollutant models for multiple analysis only when the absolute value of Pearson's correlation coefficient was < 0.3.

[b]Cox regression models were adjusted for age, gender, essential hypertension, chronic cystitis, smoking-related diagnoses, alcohol use disorders, morbid obesity, spinal cord injury, chronic liver disease, diabetes mellitus, gout, chronic kidney disease, pesticide exposures, dyslipidemia, lag0-2, season, and ambient temperature, controlled pollutants (weak correlation with THC or NMHC).

‡$p < 0.001$.
